# A year of genomic surveillance reveals how the SARS-CoV-2 pandemic unfolded in Africa

**DOI:** 10.1101/2021.05.12.21257080

**Authors:** Eduan Wilkinson, Marta Giovanetti, Houriiyah Tegally, James E. San, Richard Lessels, Diego Cuadros, Darren P. Martin, Abdel-Rahman N. Zekri, Abdoul K. Sangare, Abdoul-Salam Ouedraogo, Abdul K. Sesay, Adnène Hammami, Adrienne A. Amuri, Ahmad Sayed, Ahmed Rebai, Aida Elargoubi, Alexander J. Trotter, Alpha K. Keita, Amadou A. Sall, Amadou Kone, Amal Souissi, Ana V. Gutierrez, Andrew J. Page, Arash Iranzadeh, Arnold Lambisia, Augustina Sylverken, Azeddine Ibrahimi, Beatrice Dhaala, Bourema Kouriba, Bronwyn Kleinhans, Cara Brook, Carolyn Williamson, Catherine B. Pratt, Chantal G. Akoua-Koffi, Charles N. Agoti, Collins M. Morang’a, D. James Nokes, Daniel J. Bridges, Daniel L. Bugembe, David Baker, Deelan Doolabh, Deogratius Ssemwanga, Derek Tshabuila, Diarra Bassirou, Dominic S.Y. Amuzu, Dominique Goedhals, Dorcas Maruapula, Ebenezer Foster-Nyarko, Eddy K. Lusamaki, Edgar Simulundu, Edidah Moraa, Edith N. Ngabana, Elmostafa El Fahime, Emerald Jacob, Emmanuel Lokilo, Enatha Mukantwari, Essia Belarbi, Etienne Simon-Loriere, Etilé A. Anoh, Fabian Leendertz, Faida Ajili, Fares Wasfi, Faustinos T. Takawira, Fawzi Derrar, Feriel Bouzid, Francisca M. Muyembe, Frank Tanser, Gabriel K. Mbunsu, Gaetan Thilliez, Gemma Kay, George Githinji, Gert van Zyl, Gordon A. Awandare, Grit Schubert, Gugu P. Maphalala, Hafaliana C. Ranaivoson, Hajar Lemriss, Haruka Abe, Hela H. Karray, Hellen Nansumba, Hesham A. Elgahzaly, Hlanai Gumbo, Ibtihel Smeti, Ikhlas B. Ayed, Ilhem Boutiba-Ben Boubaker, Imed Gaaloul, Inbal Gazy, Isaac Ssewanyana, Jean B. Lekana-Douk, Jean-Claude C. Makangara, Jean-Jacques M. Tamfum, Jean-Michel Heraud, Jeffrey G. Shaffer, Jennifer Giandhari, Jingjing Li, Jiro Yasuda, Joana Q. Mends, Jocelyn Kiconco, John Morobe, John N. Nkengasong, John O. Gyapong, John T. Kayiwa, Johnathan A. Edwards, Jones Gyamfi, Jouali Farah, Joyce M. Ngoi, Joyce Namulondo, Julia C. Andeko, Julius J. Lutwama, Justin O’Grady, Kefentse A. Tumedi, Khadija M. Said, Kim Hae-Young, Kwabena O. Duedu, Lahcen Belyamani, Lavanya Singh, Leonardo de O. Martins, Madisa Mine, Magalutcheemee Ramuth, Maha Mastouri, Mahjoub Aouni, Mahmoud el Hefnawi, Maitshwarelo I. Matsheka, Malebogo Kebabonye, Manel Turki, Martin M. Nyaga, Mathabo Mareka, Matoke Damaris, Matthew Cotten, Maureen W. Mburu, Maximillian Mpina, Michael Owusu, Michael R. Wiley, Mohamed A. Ali, Mohamed Abouelhoda, Mohamed G. Seadawy, Mohamed K. Khalifa, Mooko Sekhele, Mouna Ouadghiri, Mulenga Mwenda, Mushal Allam, My V.T. Phan, Nabil Abid, Nadia Touil, Najla Kharrat, Nalia Ismael, Nedio Mabunda, Nei-yuan Hsiao, Nelson B. Silochi, Ngonda Saasa, Nicola Mulder, Patrice Combe, Patrick Semanda, Paul E. Oluniyi, Paulo Arnaldo, Peter K. Quashie, Philip A. Bester, Philippe Dussart, Placide K. Mbala, Pontiano Kaleebu, Reuben Ayivor-Djanie, Richard Njouom, Richard O. Phillips, Richmond Gorman, Robert A. Kingsley, Rosina A.A. Carr, Saâd El Kabbaj, Saba Gargouri, Saber Masmoudi, Samar Kassim, Sameh Trabelsi, Sami Kammoun, Sanaâ Lemriss, Sara H. Agwa, Sébastien Calvignac-Spencer, Seydou Doumbia, Sheila M. Mandanda, Sherihane Aryeetey, Shymaa S. Ahmed, Sikhulile Moyo, Simani Gaseitsiwe, Sonia Lekana-Douki, Sophie Prosolek, Soumeya Ouangraoua, Steve A. Mundeke, Steven Rudder, Sumir Panji, Sureshnee Pillay, Susan Engelbrecht, Susan Nabadda, Sylvie Behillil, Sylvie L. Budiaki, Sylvie van der Werf, Tapfumanei Mashe, Tarik Aanniz, Thabo Mohale, Thanh Le-Viet, Tobias Schindler, Ugochukwu J. Anyaneji, Upasana Ramphal, Vagner Fonseca, Vincent Enouf, Vivianne Gorova, Wael H. Roshdy, William K. Ampofo, Wolfgang Preiser, Wonderful T. Choga, Yaw Bediako, Yenew K Tebeje, Yeshnee Naidoo, Zaydah R. de Laurent, Sofonias K. Tessema, Tulio de Oliveira

## Abstract

The progression of the SARS-CoV-2 pandemic in Africa has so far been heterogeneous and the full impact is not yet well understood. Here, we describe the genomic epidemiology using a dataset of 8746 genomes from 33 African countries and two overseas territories. We show that the epidemics in most countries were initiated by importations, predominantly from Europe, which diminished following the early introduction of international travel restrictions. As the pandemic progressed, ongoing transmission in many countries and increasing mobility led to the emergence and spread within the continent of many variants of concern and interest, such as B.1.351, B.1.525, A.23.1 and C.1.1. Although distorted by low sampling numbers and blind-spots, the findings highlight that Africa must not be left behind in the global pandemic response, otherwise it could become a breeding ground for new variants.

## Main Text

Severe acute respiratory syndrome-related coronavirus 2 (SARS-CoV-2) emerged in late 2019 in Wuhan, China (*1, 2*). Since then, the virus has spread to all corners of the world, causing almost 150 million cases of coronavirus disease 2019 (COVID-19) and over three million deaths by the end of April 2021. Throughout the pandemic, it has been noted that Africa accounts for a relatively low proportion of reported cases and deaths – by the end of April 2021, there had been ∼4.5 million cases and ∼120 000 deaths on the continent, corresponding to less than 4% of the global burden. However, emerging data from seroprevalence surveys and autopsy studies in some African countries suggests that the true number of infections and deaths may be several fold higher than reported (*3, 4*). In addition, a recent analysis has shown that the second wave of the pandemic was more severe than the first wave in many African countries (*5*).

The first cases of COVID-19 on the African continent were reported in Nigeria, Egypt and South Africa between mid-February and early March 2020, and most countries had reported cases by the end of March 2020 (*6, 7, 8*). These early cases were concentrated amongst air travellers returning from regions of the world with high levels of community transmission. Many African countries introduced early public health and social measures (PHSM), including international travel controls, quarantine for returning travellers, and internal lockdown measures to limit the spread of the virus and give health services time to prepare (*9, 5*). The initial phase of the epidemic was then heterogeneous with relatively high case numbers reported in North Africa and Southern Africa, and fewer cases reported in other regions.

From the onset of the pandemic, genomic surveillance has been at the forefront of the COVID-19 response in Africa (*10*). Rapid implementation of SARS-CoV-2 sequencing by various laboratories in Africa enabled genomic data to be generated and shared from the early imported cases. In Nigeria, the first genome sequence was released just three days after the announcement of the first case (*6*). Similarly, in Uganda, a sequencing programme was set up rapidly to facilitate virus tracing, and the collection of samples for sequencing began immediately upon confirmation of the first case (*11*). In South Africa, the network for genomic surveillance in South Africa (NGS-SA) was established in March 2020 and within weeks genomic analysis was helping to characterize outbreaks and community transmission (*12*).

Genomic surveillance has also been critical for monitoring ongoing SARS-CoV-2 evolution and detection of new SARS-CoV-2 variants in Africa. Intensified sampling by NGS-SA in the Eastern Cape Province of South Africa in November 2020, in response to a rapid resurgence of cases, led to the detection of B.1.351 (501Y.V2) (*13*). This variant was subsequently designated a variant of concern (VOC) by the World Health Organization (WHO), due to evidence of increased transmissibility(*14*) and resistance to neutralizing antibodies elicited by natural infection and vaccines (*15*–*17*).

Here, we perform phylogenetic and phylogeographic analysis of SARS-CoV-2 genomic data from 33 African countries and two overseas territories to help characterize the dynamics of the pandemic in Africa. We show that the early introductions were predominantly from Europe, but that as the pandemic progressed there was increasing spread between African countries. We also describe the emergence and spread of a number of key SARS-CoV-2 variants in Africa, and highlight how the spread of B.1.351 (501Y.V2) and other variants contributed to the more severe second wave of the pandemic in many countries.

## Results

### SARS-CoV-2 genomic data

By 5 May 2021, 14,504 SARS-CoV-2 genomes had been submitted to the GISAID database from 38 African countries and two overseas territories (Mayotte and Réunion) (Fig. 1A). Overall, this corresponds to approximately one sequence per ∼300 reported cases. Almost half of the sequences were from South Africa (n=5362), consistent with it being responsible for almost half of the reported cases in Africa. Overall, the number of sequences correlates closely with the number of reported cases per country (Fig. 1B). The countries/territories with the highest coverage of sequencing (defined as genomes per reported case) are Kenya (n=856, one sequence per ∼203 cases), Mayotte (n=721; one sequence per ∼21 cases), and Nigeria (n=660, one sequence per ∼250 cases). Although genomic surveillance started early in many countries, few have evidence of consistent sampling across the whole year. Half of all African genomes were deposited in the first ten weeks of 2021, suggesting intensified surveillance in the second wave following the detection of B.1.351/501Y.V2 and other variants (Fig. 1C and 1D).

**Figure 1:**
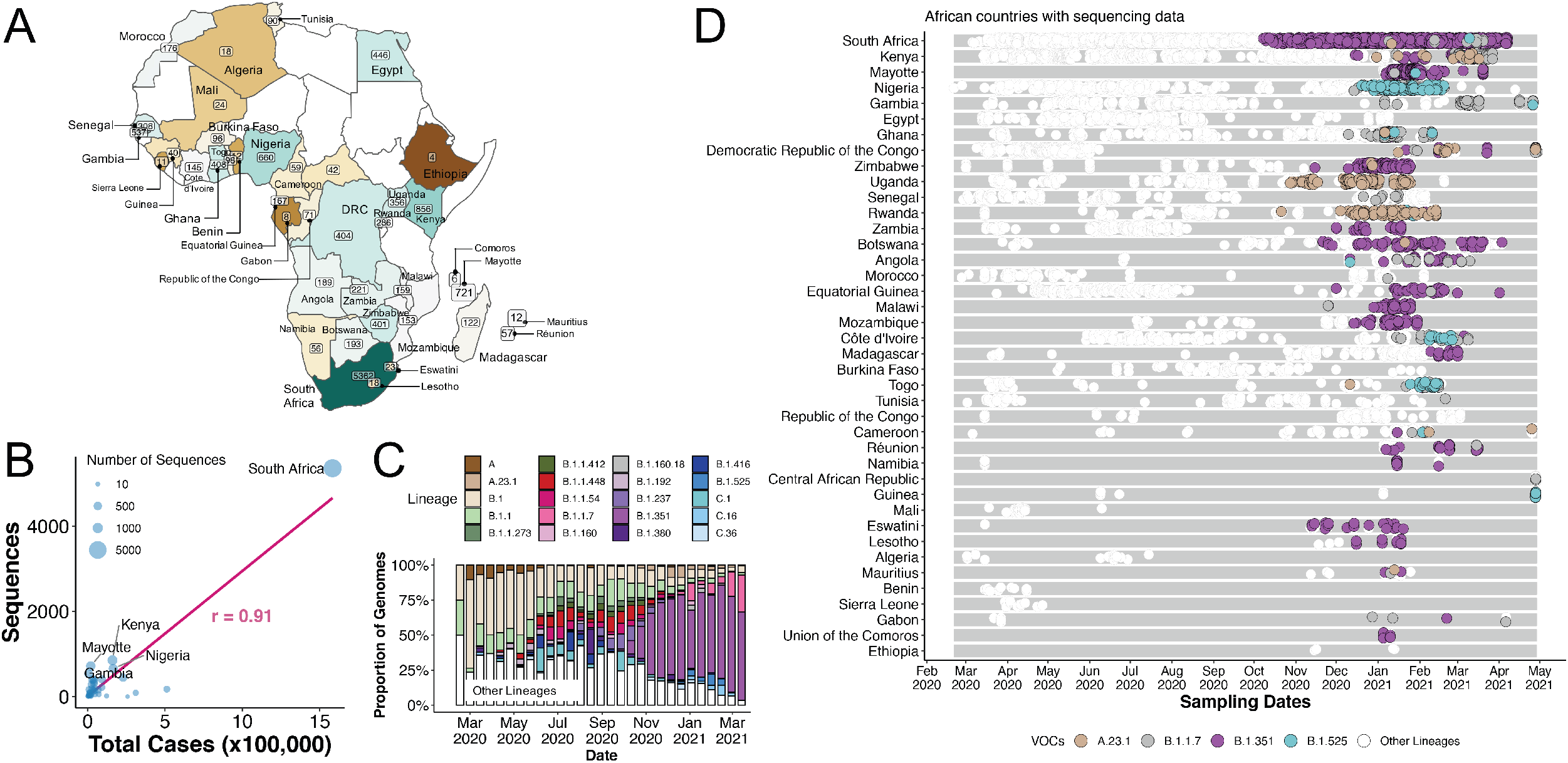
SARS-CoV-2 sequences in Africa. (A) Map of the African continent with the number of SARS-CoV-2 sequences reflected in GISAID as of 5 May 2021. (B) Regression plot of the number of viral sequences vs. the number of reported COVID-19 cases in various African countries as of 5 May 2021. Countries with >500 sequences are labelled. (C) Progressive distribution of the top 20 PANGO lineages on the African continent. (D) Temporal sampling of SARS-CoV-2 sequences in African countries (ordered by total number of sequences) through time with VOCs of note highlighted and annotated according to their PANGO lineage assignment.

### Genetic diversity and lineage dynamics in Africa

Of the 10,326 genomes retrieved from GISAID by the end of March 2021, 8,746 genomes passed quality control (QC) and met the minimum metadata requirements. These genomes from Africa were compared in a phylogenetic framework with 11,891 representative genomes from around the world. Ancestral location state reconstruction of the dated phylogeny (hereafter referred to as discrete phylogeographic reconstruction) allowed us to infer the number of viral imports and exports between Africa and the rest of the world, and between individual African countries. African genomes in this study spanned the whole global genetic diversity of SARS-CoV-2, a pattern that largely reflects multiple introductions over time from the rest of the world (Fig. 2A).

**Figure 2:**
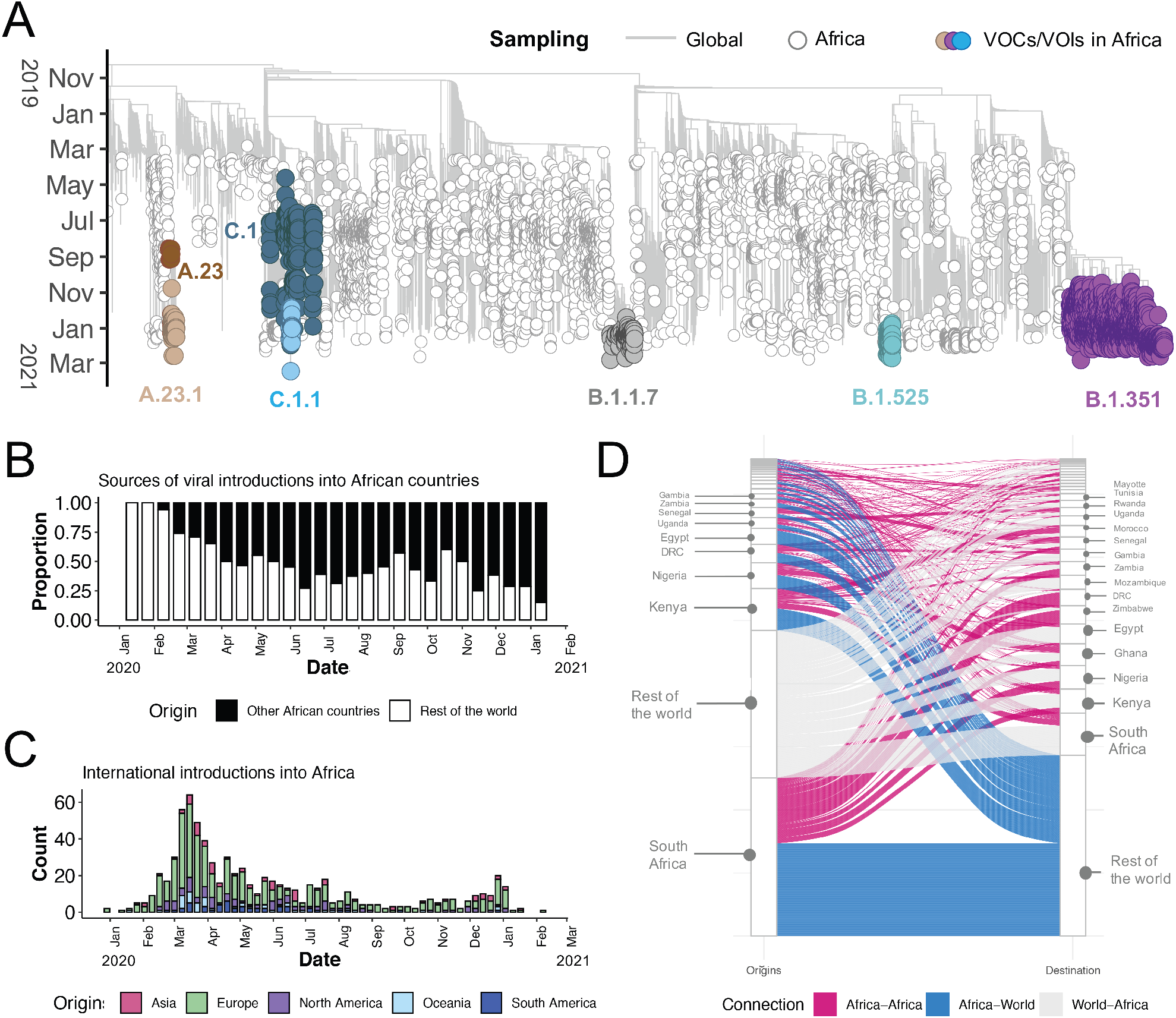
(2A) Time resolved Maximum Likelihood tree containing 8,746 high quality African SARS-CoV-2 near-full-genome sequences analyzed against a backdrop of global reference sequences. Variants of interest (VOI) and concern (VOC) are highlighted on the phylogeny. (2B) Sources of viral introductions into African countries characterized as external introductions from the rest of the world vs internal introductions from other African countries. (2C) Total external viral introductions over time into Africa. (2D) The number of viral imports and exports into and out of various African countries depicted as internal (between African countries in pink) or external (between African and non-African countries in blue and grey).

In total, we detected at least 730 viral introductions into African countries between February 2020 and February 2021, over half of which occurred before the end of May 2020. Whilst the early phase of the pandemic was dominated by importations from outside Africa, predominantly from Europe, there was then a shift in the dynamics, with an increasing number of importations from other African countries as the pandemic progressed (Fig. 2B and 2C).

South Africa, Kenya and Nigeria appear as major sources of importations into other African countries (Fig. 2D), although this is likely to be influenced by these three countries having the greatest number of deposited sequences. Particularly striking is the southern African region, where South Africa is the source for a large proportion (∼80%) of the importations to other countries in the region. The North African region demonstrates a different pattern to the rest of the continent, with more viral introductions from Europe and Asia (particularly the Middle East) than from other African countries (Fig. S2).

Africa has also contributed to the international spread of the virus with at least 356 exportation events from Africa to the rest of the world detected in this dataset. Consistent with the source of importations, most exports were to Europe (41%), Asia (26%) and North America (14%). Compared to the importation events, exportation events were more evenly distributed over time (Fig. S1). However, an increase in the number of exportation events occurred between December 2020 and March 2021, which coincided with the second wave of infections in Africa and with some relaxations of travel restrictions around the world.

The early phase of the pandemic was characterized by the predominance of lineage B.1. This was introduced multiple times to African countries and has been detected in all but one of the countries included in this analysis. After its emergence in South Africa, B.1.351 became the most frequently detected SARS-CoV-2 lineage found in Africa (n=1,769; ∼20%) (Fig. 1C). It was first sampled on 8 October 2020 in South Africa (*13*) and has since spread to 20 other African countries.

As air travel came to an almost complete halt in March/April 2020, the number(s) of detectable viral imports into Africa decreased and the pandemic entered a phase that was characterized in sub-Saharan Africa by sustained low levels of within-country movements and occasional international viral movements between neighboring countries; presumably via road and rail links between these. Though some border posts between countries were closed during the initial lockdown period (Table S1), others remained open to allow trade to continue. Regional trade in southern Africa was only slightly impacted by lockdown restrictions and quickly rebounded to pre-pandemic levels (Fig. S8) following the relaxation of restrictions between June 2020 and December 2020.

Although lineage A viruses were imported into several African countries, they only account for 1.3% of genomes sampled in Africa. Lineage A is the oldest SARS-CoV-2 lineage, representing the original Wuhan isolates of the virus from December 2019 (*18*). Despite lineage A viruses initially causing many localized clustered outbreaks, each the result of independent introductions to several countries (e.g. Burkina Faso, Cote d’Ivoire and Nigeria), they were later largely replaced by lineage B viruses as the pandemic evolved. This is possibly due to the increased transmissibility of B lineage viruses by virtue of the D614G mutation in spike (*19, 20*). However, there is evidence of an increasing prevalence of lineage A viruses in some African countries (*11*). In particular, A.23.1 emerged in East Africa and appears to be increasing rapidly in prevalence in Uganda and Rwanda (*11*). Furthermore, a highly divergent variant from lineage A was recently identified in Angola from individuals arriving from Tanzania (*21*).

### Emergence and spread of new SARS-CoV-2 variants

In order to determine how some of the key SARS-CoV-2 variants are spreading within Africa, we performed phylogeographic analyses on the VOC B.1.351, the VOI B.1.525, and on two additional variants that emerged and that we designated as VOIs for this analysis (A.23.1 and C.1.1). These African VOCs and VOIs have multiple mutations on Spike glycoprotein and molecular clock analysis of these four datasets provided strong evidence that these four lineages are evolving in a clocklike manner (Fig. 3A, 3B).

**Figure 3:**
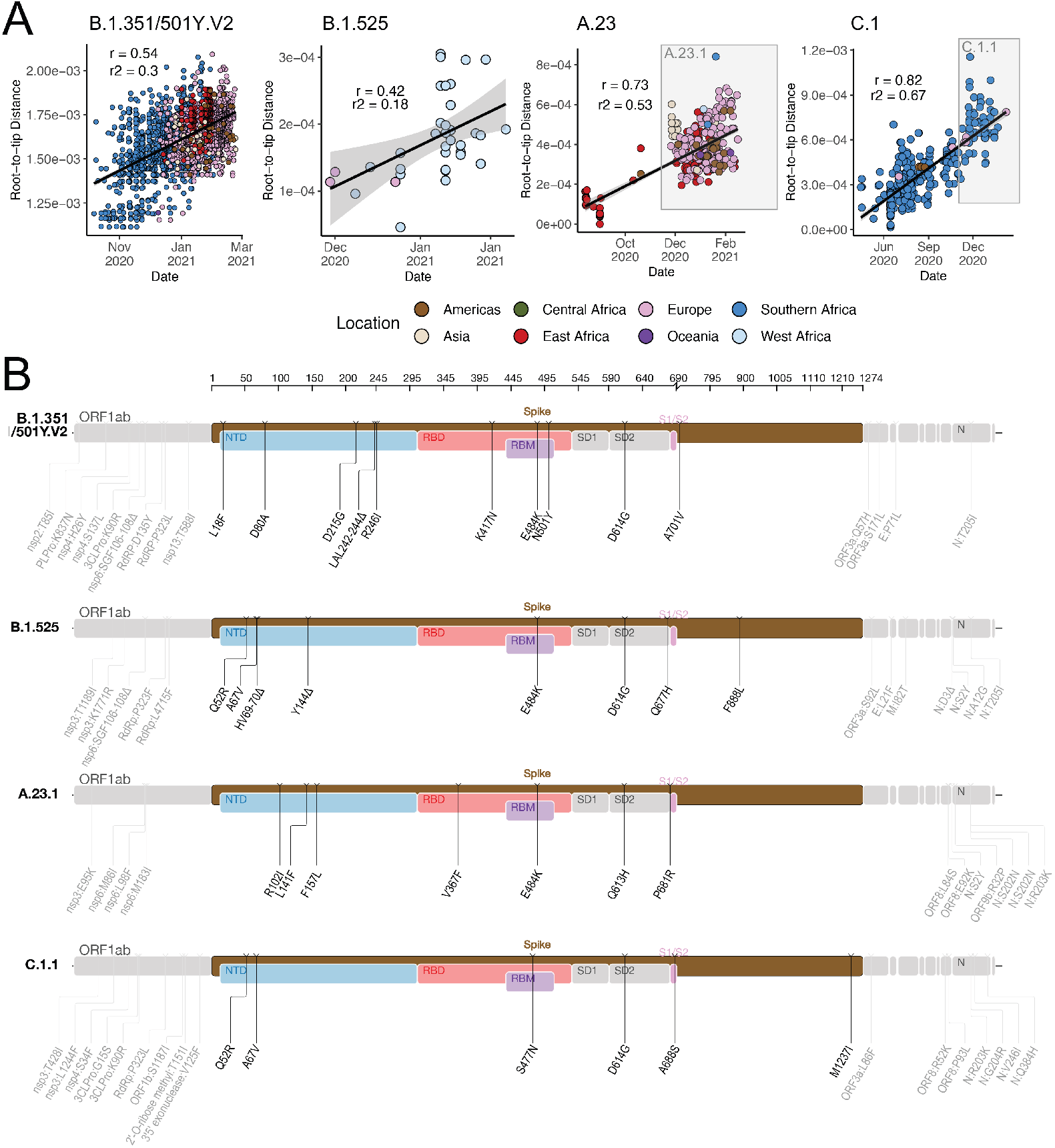
(A) Root-to-tip regression plots for four lineages of interest. C.1 and A.23 show continued evolution into VOIs C.1.1 and A.23.1 respectively. (B) Genome maps of four VOCs/VOIs where the spike region is shown in detail and in color and the rest of the genome is shown in grey.

B.1.351 was first sampled in South Africa in October 2020, but phylogeographic analysis suggests that it emerged earlier, around August 2020. It is defined by ten mutations in the spike protein, including K417N, E484K and N501Y in the receptor-binding domain (Fig. 3B). Following its emergence in the Eastern Cape, it spread extensively within South Africa (Fig. 4A). By November 2020, the variant had spread into neighbouring Botswana and Mozambique and by December 2020 it had reached Zambia and Mayotte. Within the first three months of 2021, further exports from South Africa into Botswana, Zimbabwe, Mozambique and Zambia occurred. By March 2021, B.1.351 had become the dominant lineage within most Southern African countries as well as the overseas territories of Mayotte and Réunion (Fig. S3). Our phylogeographic reconstruction also demonstrates movement of B.1.351 into East and Central Africa directly from southern Africa. A discrete phylogeographic analysis of a wider sample of viruses suggests that the transmission to West Africa may have occurred via East Africa, with a possible European intermediary (Fig. S4).

**Figure 4:**
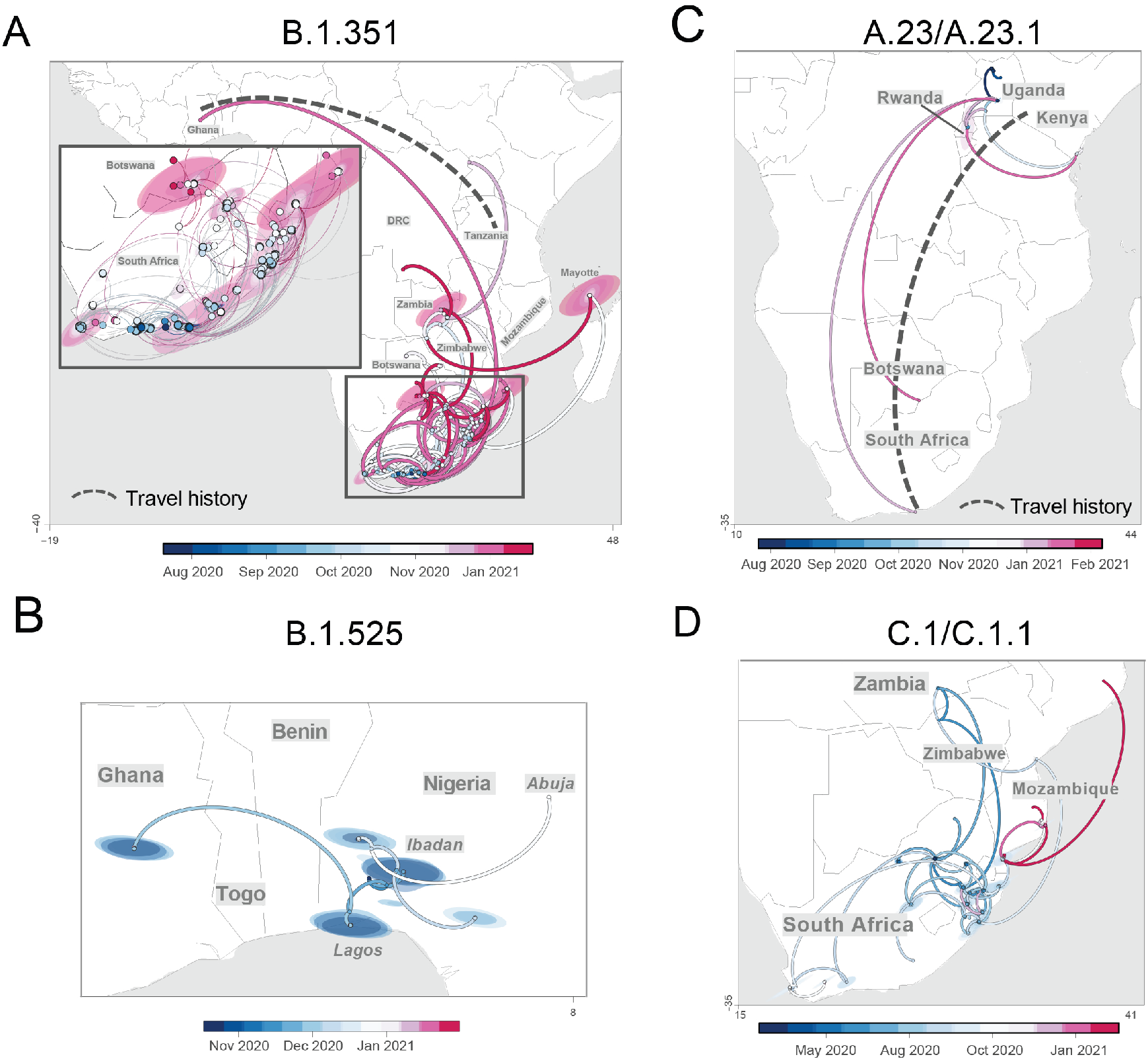
Phylogeographic reconstruction of the spread of four VOCs/VOIs across the African continent using sequences showing strict continuous transmission across geographical regions. Curved lines denote the direction of transmission in the anti-clockwise direction. Solid lines show transmission paths as inferred by phylogeographic reconstruction and colored by date, whereas dashed lines show known travel history of the particular case considered.

B.1.525 is a VOI defined by six substitutions in the spike protein (Q52R, A67V, E484K, D614G, O677H and F888L), and two deletions in the N-terminal domain (HV69-70Δ and Y144Δ). This was first sampled in the United Kingdom in mid-December 2020, but our phylogeographic reconstruction suggests that the variant originated in Nigeria in November 2020 (95% highest posterior density (HPD) 2020-11-01 to 2020-12-03) (Fig. 4B). Since then it has spread throughout much of Nigeria and neighbouring Ghana. Given sparse sampling from other neighbouring countries within West and Central Africa (Fig. 1A & 1C), the extent of the spread of this VOI in the region is not clear. Beyond Africa, this VOI has spread to Europe and the US (Fig. S4).

We designated A.23.1 and C.1.1 as VOIs for the purposes of this analysis, as they present good examples of the continued evolution of the virus within Africa(*11, 13*). Lineage A.23, characterized by three spike mutations (F157L, V367F and Q613H), was first detected in a Ugandan prison in Amuru in July 2020 (95% HPD: 2020-07-15 to 2020-08-02). From there, the lineage was transmitted to Kitgum prison, possibly facilitated by the transfer of prisoners. Subsequently, the A.23 lineage spilled into the general population and spread to Kampala, adding other spike mutations (R102I, L141F, E484K, P681R) along with additional mutations in nsp3, nsp6, ORF8 and ORF9, prompting a new lineage classification, A.23.1 (Fig. 3A & 3B). Since the emergence of A.23.1 in September 2020 (95% HPD: 2020-09-02 to 2020-09-28), it has spread regionally into neighbouring Rwanda and Kenya and has now also reached South Africa and Botswana in the south and Ghana in the west (Fig. 4C). However, our phylogeographic reconstruction of A.23.1 suggests that the introduction into Ghana may have occurred via Europe (Fig. S4), whereas the introductions into southern Africa likely occurred directly from East Africa. This is consistent with epidemiological data suggesting that the case detected in South Africa was a contact of an individual who had recently travelled to Kenya.

Lineage C.1 emerged in South Africa in March 2020 (95% HPD: 2020-03-13 to 2020-04-17) during a cluster outbreak prior to the first wave of the epidemic(*13*). C.1.1 is defined by the spike mutations S477N, A688S, M1237I and also contains the Q52R and A67V mutations similar to B.1.525 (Fig. 3B). A continuous trait phylogeographic reconstruction of the movement dynamics of these lineages suggests that C.1 emerged in the city of Johannesburg and spread within South Africa during the first wave (Fig. 4D). Independent exports of C.1 from South Africa led to regional spread to Zambia (June-July, 2020) and Mozambique (July-August 2020), and the evolution to C.1.1 seems to have occurred in Mozambique around mid-September 2020 (95% HPD: 2020-09-07 to 2020-10-05). In depth analysis of SARS-CoV-2 genotypes from Mozambique suggest that the C.1.1. lineage was the most prevalent in the country until the introduction of B.1.351, which has dominated the epidemic since (Fig. S3).

The VOC B.1.1.7, which was first sampled in Kent, England in September 2020(*22*), has also increased in prevalence in several African countries (Fig. S3) To date, this VOC has been detected in eleven African countries, as well as the Indian Ocean islands of Mauritius and Mayotte (Fig. S5). The time-resolved phylogeny suggests that this lineage was introduced into Africa on at least 16 occasions between November 2020 and February 2021 with evidence of local transmission in Nigeria and Ghana.

## Conclusions

Our phylogeographic reconstruction of past viral dissemination patterns suggests a strong epidemiological linkage between Europe and Africa, with 64% of detectable viral imports into Africa originating in Europe and 41% of detectable viral exports from Africa landing in Europe (Figure 1C). This phylogeographic analysis also suggests a changing pattern of viral diffusion into and within Africa over the course of 2020. In almost all instances the earliest introductions of SARS-CoV-2 into individual African countries were from countries outside Africa.

High rates of COVID-19 testing and consistent genomic surveillance in the south of the continent have led to the early identification of VOCs such as B.1.351 and VOIs such as C.1.1(*13*). Since the discovery of these southern African variants, several other SARS-CoV-2 VOIs have emerged in different parts of the world, including elsewhere on the African continent, such as B.1.525 in West Africa and A.23.1 in East Africa). There is strong evidence that both of these VOIs are rising in frequency in the regions where they have been detected, which suggests that they may possess higher fitness than other variants in these regions. Although more focused research on the biological properties of these VOIs is needed to confirm whether they should be considered VOCs, it would be prudent to assume the worst and focus on limiting their spread. It is quite clear that we are currently seeing the virus adapting to immune responses that were developed by people who became infected during the first and second waves of the epidemic in Africa. It will be important to investigate how these different variants compete against one another if they occupy the same region.

Our focused phylogenetic analysis of the B.1.351 lineage revealed that in the final months of 2020 this variant spread from South Africa into neighbouring countries, reaching as far north as the DRC by February 2021. This spread may have been facilitated through rail and road networks that form major transport arteries linking South Africa’s ocean ports to commercial and industrial centres in Botswana, Zimbabwe, Zambia and the southern parts of the DRC. The rapid, apparently unimpeded spread of B.1.351 into these countries suggests that current land-border controls that are intended to curb the international spread of the virus are ineffective. Perhaps targeted testing of cross-border travellers, genotyping of positive cases and the focused tracking of frequent cross-border travellers such as long distance truckers, would more effectively contain the spread of future VOCs and VOIs that emerge within this region.

The dominance of VOIs and VOCs in Africa has important implications for vaccine rollouts on the continent. For one, slow rollout of vaccines in most African countries creates an environment in which the virus can replicate and evolve: this will almost certainly produce additional VOCs, any of which could derail the global fight against COVID-19. On the other hand, with the already widespread presence of known variants, difficult decisions balancing reduced efficacy and availability of vaccines have to be made. This also highlights how crucial it is that trials are done. From a public health perspective, genomic surveillance is only one item in the toolkit of pandemic preparedness. It is important that such work is closely followed by genotype to phenotype research to determine the actual significance of continued evolution of SARS-CoV-2 and other emerging pathogens.

The rollout of vaccines across Africa has been painfully slow (Fig. S6). There have, however, been notable successes that suggest the situation is not hopeless. The small island nation of the Seychelles had vaccinated 70% of its population by May 2021. Morocco has kept pace with many developed nations and by mid-March had vaccinated ∼16% of its population. Rwanda, one of Africa’s most resource constrained countries, had, within three weeks of obtaining its first vaccine doses in early March, managed to provide first doses to ∼2.5% of its population. For all other African countries, at the time of writing, vaccine coverage (first dose) was <1.0% of the general population.

The effectiveness of molecular surveillance as a tool for monitoring pandemics is largely dependent on continuous and consistent sampling through time, rapid virus genome sequencing and rapid reporting. When this is achieved, molecular surveillance can ensure the early detection of changing pandemic characteristics. Further, when such changes are discovered, molecular surveillance data can also guide public health responses. In this regard, the molecular surveillance data that are being gathered by most African countries are less useful than they could be. For example, the time-lag between when virus samples are taken and when sequences for these samples are deposited in sequence repositories is so great in some cases that the primary utility of genomic surveillance data is lost (Fig. S9). More recent sampling and prompt reporting is crucial to reveal the genetic characteristics of currently circulating viruses in these countries.

The patchiness of African genomic surveillance data is therefore the main weakness of our study. However, there is evidence that the situation is improving, with ∼50% of African SARS-CoV-2 genome sequences having been submitted to the GISAID database within the first 10-weeks of 2021. While the precise factors underlying this surge in sequencing effort are unclear, important drivers are almost certainly both increased global interest in genomic surveillance following the discovery of multiple VOCs and VOIs since December 2020. We cannot reject that the observed increase in exports from Africa may be due to intensified sequencing activity following the detection of variants around the world. It is important to note here that phylogeographic reconstruction of viral spread is highly dependent on sampling where there is the caveat that the exact routes of viral movements between countries cannot be inferred if there is no sampling in connecting countries. Furthermore, our efforts to reconstruct the movement dynamics of SARS-CoV-2 across the continent are almost certainly biased by uneven sampling between different African countries. It is not a coincidence that we identified South Africa, Kenya and Nigeria, which have sampled and sequenced the most SARS-CoV-2 genomes, as major sources of viral transmissions between sub-Saharan African countries. However, these countries had also the highest number of infections, which may decrease the sampling biases (Fig. 1A).

The reliability of genomic surveillance as a tool to prevent the emergence and spread of dangerous variants is dependent on the intensity with which it is embraced by national public health programs. As with most other parts of the world, the success of genomic surveillance in Africa requires more samples being tested for COVID-19, higher proportions of positive samples being sequenced within days of sampling, and persistent analyses of these sequences for concerning signals such as (*i*) the presence of novel non-synonymous mutations at genomic sites associated with pathogenicity and immunogenicity, (*ii*) evidence of positive selection at codon sites where non-synonymous mutations are observed, and (*iii*) evidence of lineage expansions. In spite of limited sampling, Africa has identified many of the VOCs and VOIs that are being transmitted across the world. Detailed characterization of the variants and their impact on vaccine induced immunity is of extreme importance. If the pandemic is not controlled in Africa, we may see the production of vaccine escape variants that may profoundly affect the population in Africa and across the world.

## Data Availability

All genomes were available at GISAID. Short reads availabe at short read archive (SRA) of the NCBI/EBI

## Acknowledgments

We wish to acknowledge the contribution of Lynn Tyers, Kruger Maria and Innocent Mudau from the National Genomics Surveillance of South Africa (NGS-SA) platform for their contribution towards the sequencing effort in Cape Town; South Africa. Similarly, we wish to thank Aya M Elsaame, Shimaa M, Elsayed and Reham M. Darwish from the Faculty of Medicine Ain Shams Research institute (MASRI), for their efforts towards sequencing in Egypt. Sidy Bane, Moumine Sanogo, Dramane Diallo, Antieme Combo Georges Togo and Aminatou Coulibaly from the University Clinical Research Centre (UCRC), at the University of Sciences, Techniques and Technologies of Bamako we wish to extend our thanks for the contribution they have made towards sequencing efforts in Mali.

## Author contributions

**Conceptualization:** EW, HT, RL, TdO, KOD, RAD, JG, JOG

**Methodology:** EW, MG, DT, JES, KOD, RAAC, RAD, JG

**Investigation:** EW, MG, JES, HT, SE,CW, NA, AS, MM, KOD, RAAC, JG, DG, MMN, PAB

**Sampling:** MR

**Sequencing:** JG, LS, YN, DT, SP, SE, †BK, TM, HG, CMM, CW MC, DLB, MVTP, PK

**Visualization:** JES, HT

**Funding acquisition:** TdO, MC, AR, KOD, RAK

**Project administration:** AR, KOD

**Supervision:** TdO, RL, KOD, JOG, RAK

**Writing – original draft:** EW, JES, HT, DPM

**Writing – review & editing:** DPM, EW, CW, MC, RAK

## Competing interests

### Data and materials availability

All input files (e.g. alignments or XML files), all resulting output files and scripts used in the study are shared publicly on GitHub (https://github.com/krisp-kwazulu-natal/africa-covid19-genomics).

### Supplementary Figures and Tables

**Supplementary Figure S1:** *Number of importation and exportation events for various subregions on the African continent. African subregions are defined based on the African Union classification scheme*.

**Supplementary Figure S2:** *Numbers of exportation events from the African continent to the rest of the world*.

**Supplementary Figure S3:** *PANGO lineages through time for a select number of African countries*.

**Supplementary Figure S4:** *Maximum clade credibility phylogeographic trees including all global VOC or VOI samples. Branch colours represent most probable inferred locations of ancestral viruses. Numbers at internal nodes represent clade posterior probabilities*.

**Supplementary Figure S5:** *Time scaled phylogeny of the B*.*1*.*1*.*7 lineage. This phylogenetic cluster was extracted from the large dated phylogeny in Figure 2A. African sequences are highlighted by large circles, while non-African sequences appear as smaller dots. The branches are scaled in calendar time*.

**Supplementary Figure S6: Epidemiological metricies of COVID-19 on the African continent**. *Clockwise from top left: reported COVID-19 cases per million individuals; reported COVID-19 attributed mortalities per million individuals; numbers of COVID-19 tests performed per 1,000 individuals; and numbers vaccinated per 100 individuals*.

**Supplementary Figure S7:** *Epidemiological heatmaps of cases and deaths for various subregions on the Afican continent. African subregions are defined based on the African Union classification scheme*.

**Supplementary Figure S8**. *Total monthly international trade values in US million dollars in 2020 for A) exported goods from South Africa; and B) imported goods to South Africa with the following neighbouring countries: Botswana, Democratic Republic of the Congo, Eswatini, Lesotho, Malawi, Mozambique, Namibia, Zambia, and Zimbabwe. Source: UN Comtrade Database*.

**Supplementary Figure S9:** *Graph of days from sampling to submission in various African countries*.

**Supplementary Table S1**. *Status and restrictions of land border posts in South Africa as of Feb 19, 2021*.

**Supplementary Table S2:** *Variants of Concern/Note (VoC/Ns) in Africa*

**Supplementary Table S3:** *Sampling or surveillance strategies in various participating institutions*.

## Materials and Methods

### Data quality control

10,326 African complete and near-complete genome sequences were retrieved from GISAID on 16 March 2021 (2pm SAST). Sampling strategies in various participating countries are outlined in Supplementary Table S3. Prior to phylogenetic reconstruction we removed low quality sequences, which included those identified as being of low quality by NextClade (n=18; https://clades.nextstrain.org), those with missing sampling dates (n = 189), those with <90% coverage (n = 1,017), those with > 40 SNPs (n = 39), those with >10 ambiguous base-calls per genome (n = 128), and those with clustered SNPs (n = 189).

High quality African near-complete genome sequences (n=8,746) were aligned against an extensive reference dataset of 11,891 SARS-CoV-2 sequences from around the world that included sequences sampled since the start of the outbreak, including all those sampled up until the end of February 2020.

### Phylogenetic reconstruction

The African sequences were aligned against the reference panel using MAFFT v7.471 (*23*). The first 100 and last 50 bases as well as positions 13402, 24389 and 24390 relative to the reference strain (Wuhan-Hu-1: Accession NC_045512) were masked. The subsequent alignment was used to infer a maximum likelihood (ML) phylogenetic tree in IQTREE v1.6.9 (*24*). The tree was inferred with the general time reversible (GTR) model of nucleotide substitution and a proportion of invariable sites (+I). To infer some confidence measures of branches in the phylogeny and for subsequent downstream analyses we performed 100 bootstrap replicates using Booster (*25*).

The raw ML tree topology was used to estimate the number of viral transmission events between various Africa countries and the rest of the world. TreeTime (*26*) was used to transform this ML tree topology into a dated tree using a constant rate of 8.0 × 10^−4^ nucleotide substitutions per site per year, after the exclusion of outlier sequences. A migration model was fitted to the resulting time-scaled phylogenetic tree in TreeTime, mapping country and regional locations to tips and internal nodes. Using the resulting annotated tree topology we could count the number of transitions between Africa and the rest of the world.

### Lineage classification

We used the dynamic lineage classification method called Phylogenetic Assignment of Named Global Outbreak LINeages (PANGOLIN) (*27*). This was aimed at identifying the most epidemiologically important lineages of SARS-CoV-2 circulating within the African continent and to identify the lineage dynamics within African regions and across the continent. For the purpose of clarity, we define a lineage as a linear chain of viruses in a phylogenetic tree showing connection from the ancestor to the most recent descendant. A unique variant refers to a genetically distinct virus with different mutations to other viruses of the same lineage. Variants of concern (VOC) and variants of interest (VOI) were designated based on the World Health Organization framework as of 13 April 2021. We included two other lineages, namely A.23.1 and C.1.1, and designated them as VOI for the purposes of this analysis. We included these two as they demonstrated continued evolution of African lineages into potentially more transmissible variants with the acquisition of mutations in the spike glycoprotein.

### Phylogeographic reconstruction

VOCs and VOIs that emerged on the African continent (B.1.351, B.1.525, A.23.1 and C.1.1) were marked on the time-resolved phylogenetic tree constructed above. Genome sequences from these four lineages were extracted for phylogeographic reconstruction. First, we investigated the dynamics of SARS-CoV-2 infection and virus lineage movements over longer distances (through Europe or East to West Africa) using a sampled set of time-scaled phylogenies and the sampling location of each geo-referenced SARS-CoV-2 sequence. We discretized sequence sampling locations by considering distinct geographic areas and/or regions (in and outside Africa) as shown in Supplementary Figure S4.

Initially, discrete phylogeographic reconstructions were conducted for all VOC and VOI using the asymmetric discrete trait model implemented in BEASTv1.10.4 (*28*). From those estimates we then modelled the phylogenetic diffusion and spread of the lineages on the African continent by analysing localized transmission (between neighboring countries) using a flexible relaxed random walk (RRW) diffusion model (*29*) that accommodates branch-specific variation in rates of dispersal with a Cauchy distribution. For each sequence, latitude and longitude coordinates were attributed to the lowest administrative level locator in GISAID.

Multiple sequence alignments were performed for each lineage with MAFFT v7.471. Maximum likelihood trees for each of the alignments were inferred in IQTREE v1.6.9 (GTR+I). Prior to phylogeographic reconstruction each cluster/lineage was assessed for molecular clock signal in TempEst v1.5.3 (*30*) following the removal of potential outliers that may violate the molecular clock assumption. Markov Chain Monte Carlo (MCMC) analyses were set up in BEAST v1.10.4 in duplicate for 100 million interactions and sampling every 10,000 steps in the chain. Convergence for each run was assessed in Tracer v1.7.1 (ESS for all relevant model parameters >200). Maximum clade credibility trees for each run were summarized using TreeAnnotator after discarding the initial 10% as burn-in. We used the R package “seraphim” (*31*) to extract and map spatiotemporal information embedded in the posterior trees. Note that a transmission link on the phylogeographic map can denote one or more transmission events depending on the phylogeographic inference.

### Epidemiological modeling

Data on regional trade of all imported and exported goods between South Africa and other Eastern and Southern African countries during 2020 was extracted from the United Nations Comtrade Database (*32*), which records trade statistics for more than 5,000 commodity groups by the Harmonized System. Data for cumulative COVID-19 cases and related deaths, vaccinated people, and cumulative numbers of COVID-19 tests performed by March 30, 2021 were obtained from the Johns Hopkins University database (*33*). Country level maps of each variable were created using ArcGIS^®^ by ESRI version 10.5 (http://www.esri.com).

