## Supplementary figures and images for "A year of genomic surveillance reveals how the SARS-CoV-2 pandemic unfolded in Africa"

### Supplemtary Figure 1

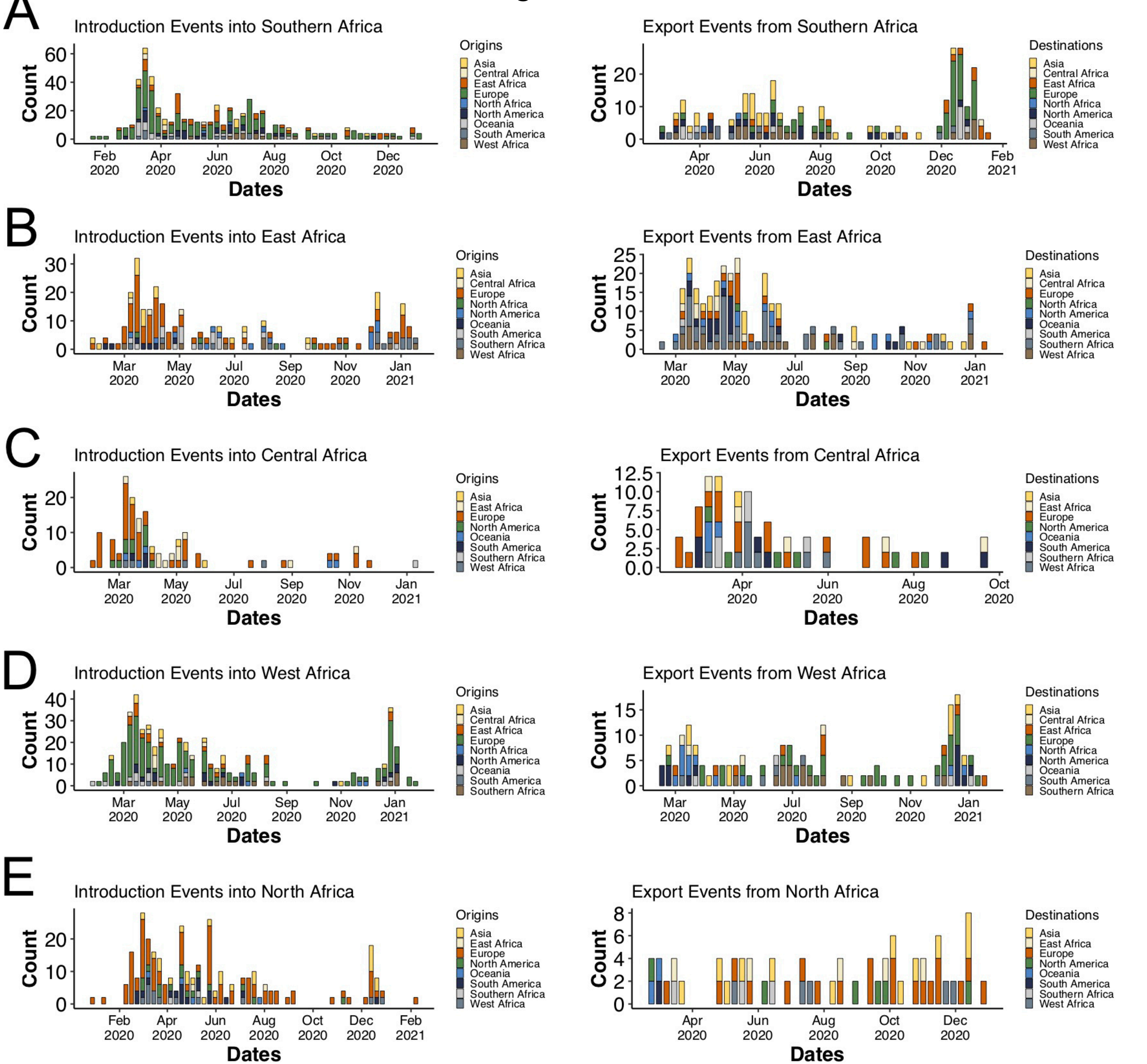

### Supplemtary Figure 2

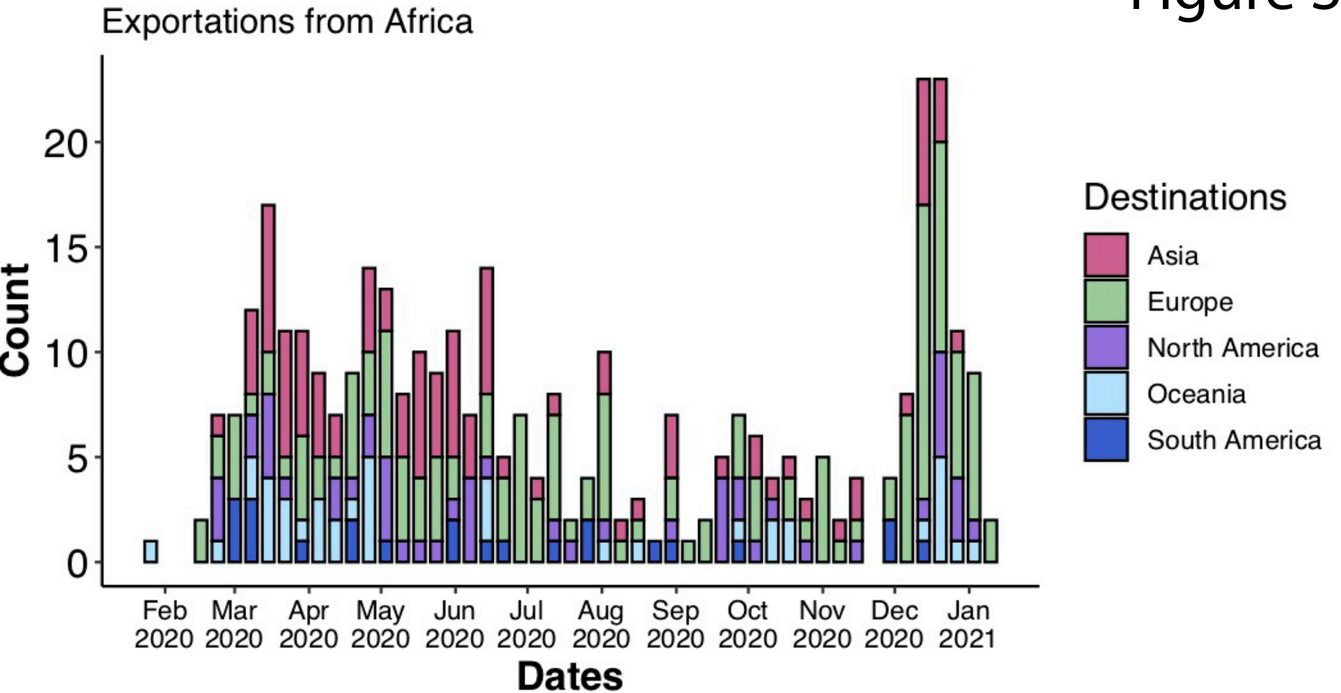

### Supplemtary Figure 3

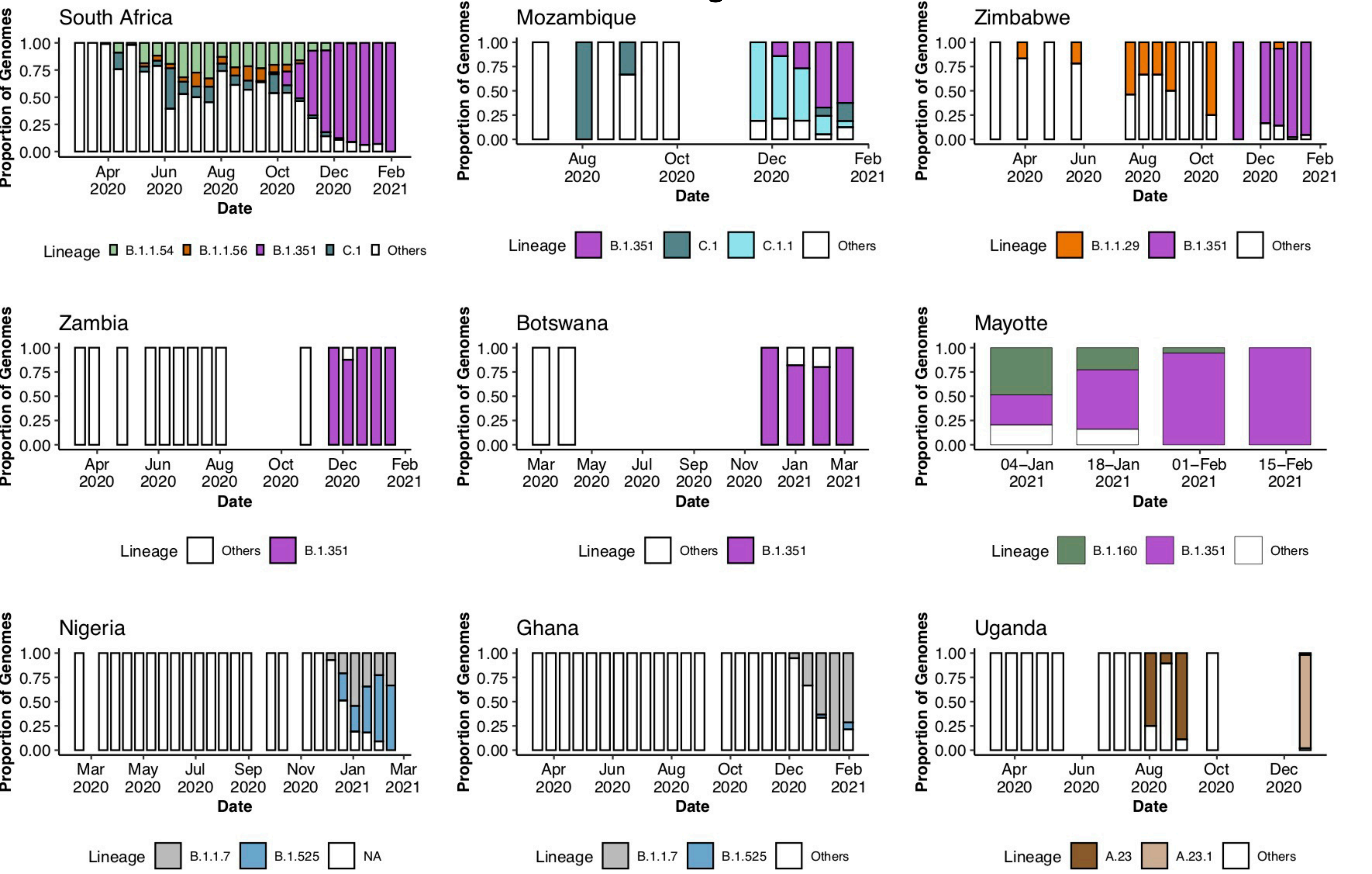

### Supplemtary Figure 4

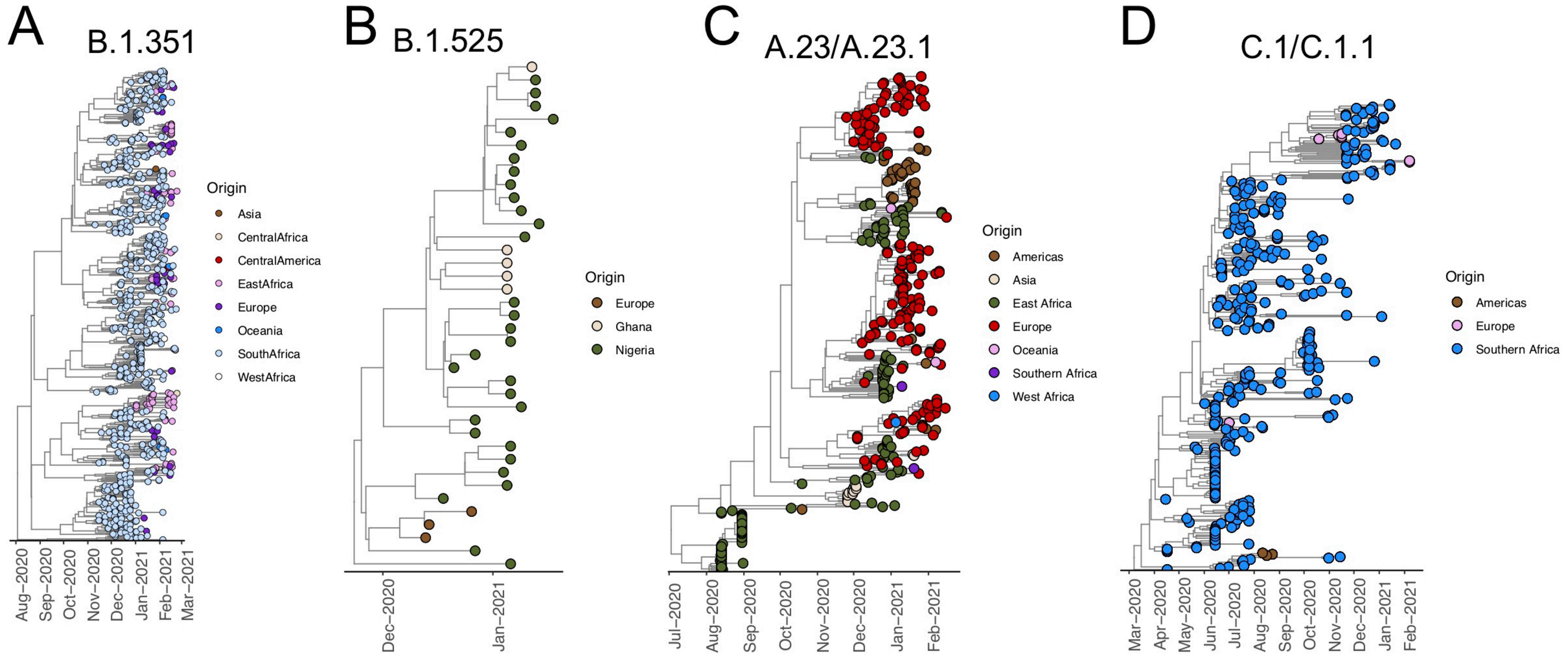

### Supplemtary Figure 5

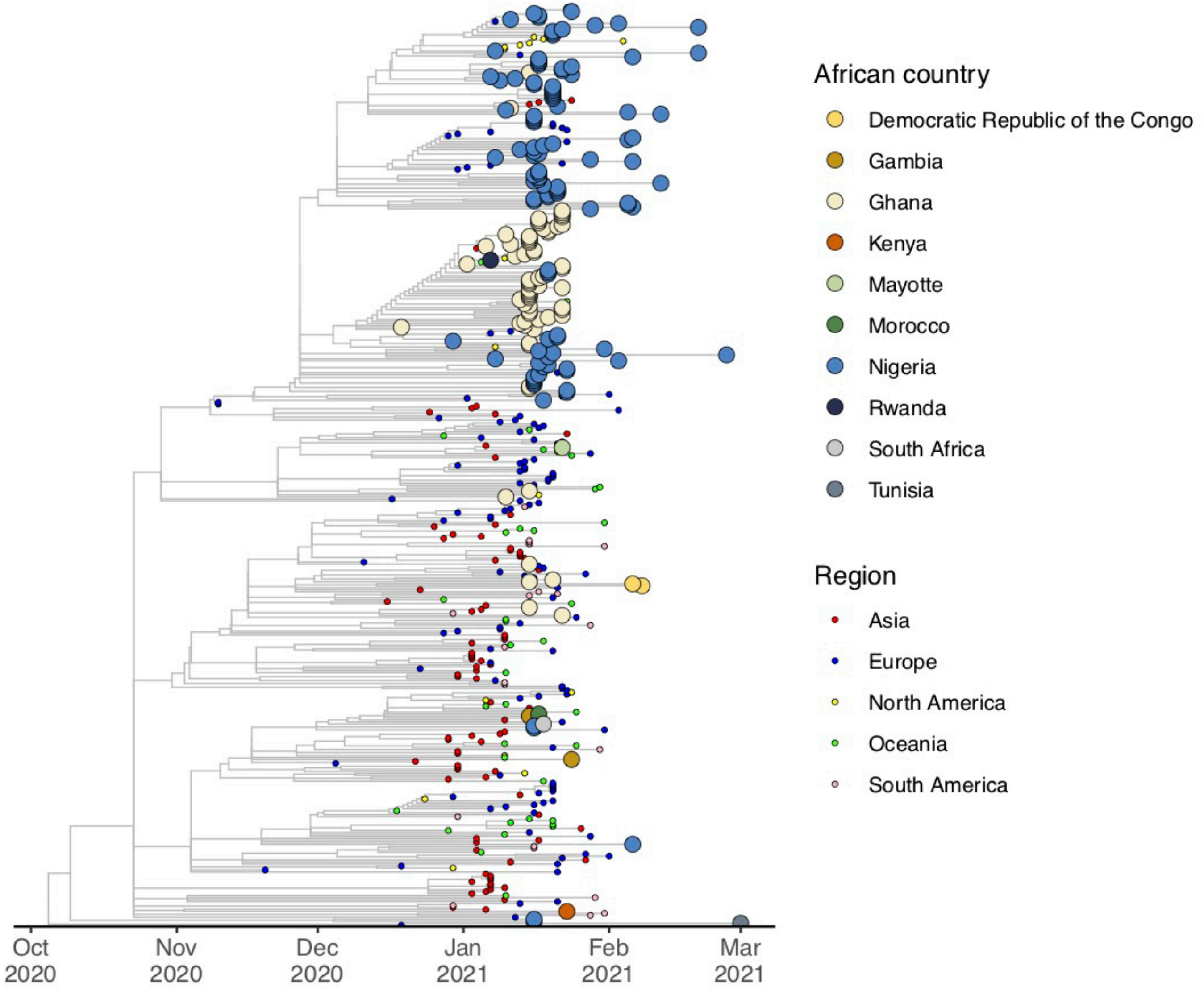

### Supplemtary Figure 6

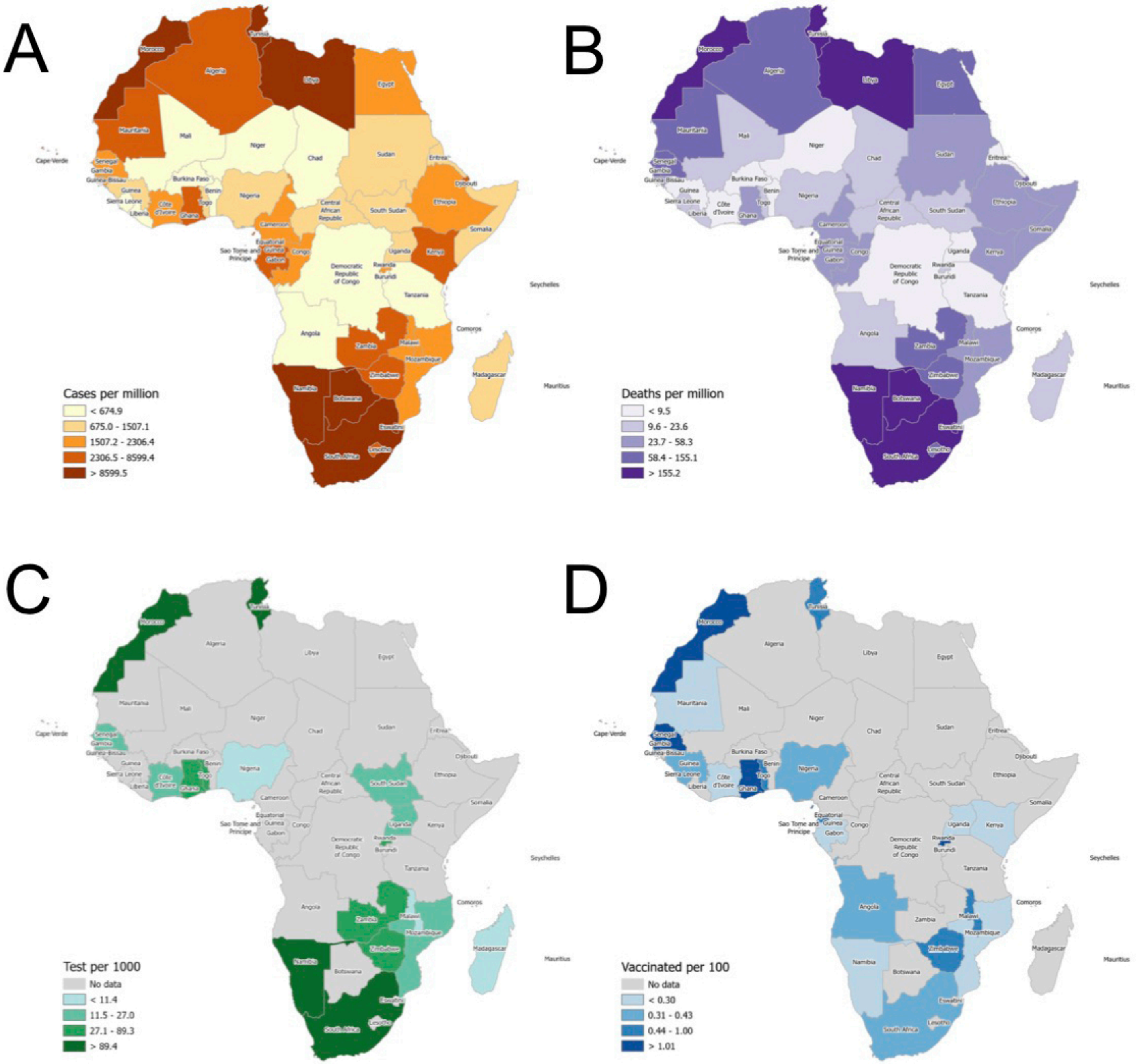

### Supplemtary Figure 7

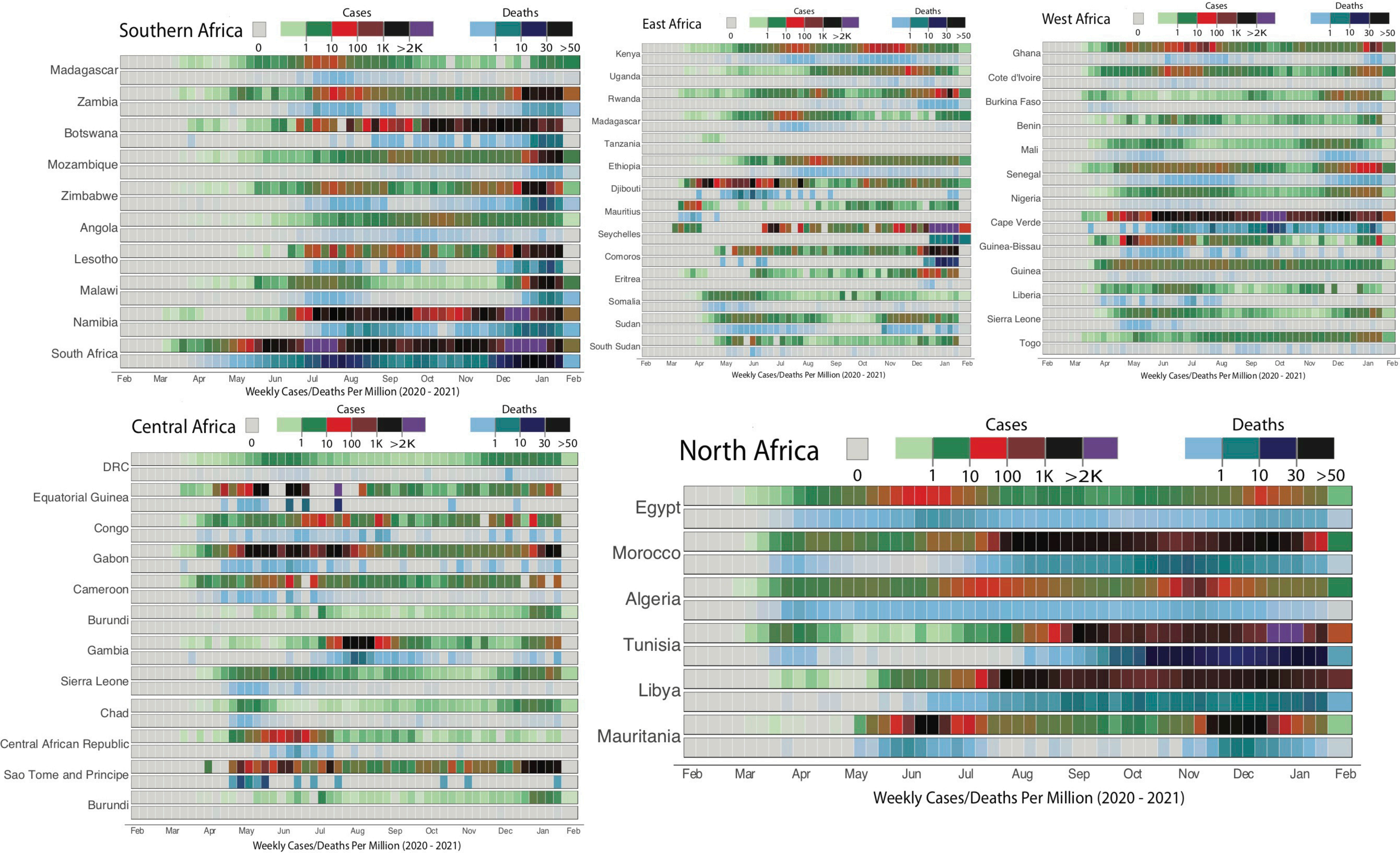

### Supplemtary Figure 8

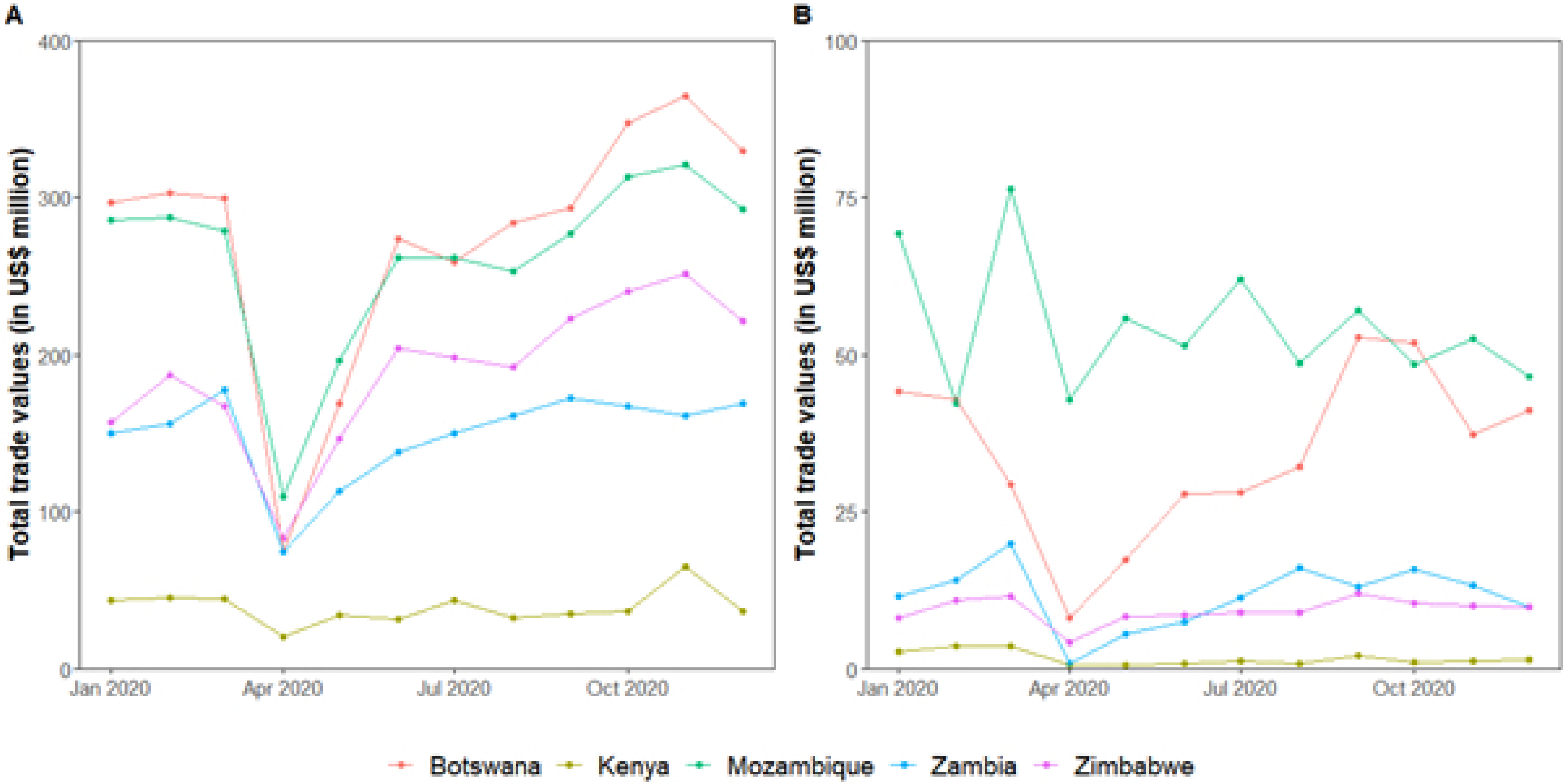
