## Supplementary Table for "A year of genomic surveillance reveals how the SARS-CoV-2 pandemic unfolded in Africa"

**Supplementary Table S1. Status and restrictions of land border posts in South Africa as of Feb 19, 2021.**

| Country route | Number of land border posts | | Restrictions |
|---|---|---|---|
| | Closed (n/N) | Open (n/N) | |
| South Africa - Botswana | 13/17 | 4/17 | · All passengers passing through the border posts are required to present a medical certificate with a negative COVID-19 test result issued within 72 hours or get tested upon arrival and subject to quarantine in a government holding facility. The entry to Zimbabwe requires a negative COVID-19 test result that is within 48 hours.<br>· Rail, ocean, air and road transport is permitted for the movement of cargo to and from other countries, subject to national legislation and any directions.<br>· All borders were closed on Jan 11, 2021 then reopened on February 15, 2021. |
| South Africa - eSwatini | 6/11 | 5/11 | |
| South Africa - Lesotho | 7/13 | 6/13 | |
| South Africa - Mozambique | 2/4 | 2/4 | |
| South Africa - Namibia | 4/6 | 2/6 | |
| South Africa - Zimbabwe | 0/1 | 1/1 | |

**Supplementary Table S2. Variants of Concern/Note (VoC/Ns) in Africa**

| **Variant Name** | **Lineage** | **Date Range** | **Spike Mutations of Biological Significance (all mutations)** | **Impact** | **Countries** |
|---|---|---|---|---|---|
| N501Y.V2 | B.1.351 | Oct. 2020 – Feb. 2021 | K417N, E484K, N501Y | Transmissibility, Escape Neutralization, ACE binding Affinity | South Africa, DRC, Mayotte, La Reunion, Zambia, Botswana, Congo, Kenya, Rwanda, |
| A.23, A.23.1 | A.23.1 | Dec. 2020 – Feb 2021 | V367F, Q613H | Infectivity | Uganda, Rwanda, Ghana, South Africa, Zambia, Botswana |
| C.1.1 | C.1. | | S477N | | Mozambique, |
| B.1.525 | B.1.525 | Dec. 2020 - Feb 2021 | E484K, Q677H, F888L | Escape Neutralization, ACE binding Affinity | Nigeria, Ghana, Mayotte, Côte d'Ivoire/Bouaké Algeria |
| A.27/N501Y.V4 | A.27 | Jan 2021 - Feb 2021 | L18F, L452R, N501Y, A653V, H655Y, Q677H, D796Y, G1219V | under investigation (VUI not VOC) | Mayotte, Europe, Ghana, Côte d'Ivoire/Bouaké |
| N501Y.V3 | | | | | Brazil |
| B.1.160 | B.1.160 | | D614G, S477N | confirmed reinfection (**under investigation**) | **Tunisia** (reinfection), Large European lineage Ghana |
| N501Y | B.1.1.7 | Jan - Mash2021 | D614G, N501Y, del69-70, | Transmissibility | **Ghana, Morocco Algeria,** Côte d'Ivoire/Bouaké, DRC |

**Supplementary Table S3. Sampling or surveillance strategies in various participating institutions.**

| Country | Proportion of cases sequenced | Sampling strategies | | | | Other (details) |
|---|---|---|---|---|---|---|
| | | Regular surveillance (random sampling) | Cluster/outbreak investigations | Surveillance of imported cases (linked to border testing) | Investigation of re-infections | |
| South Africa | 0.20% | Yes | Yes | No | Yes | Sequencing of infections in vaccine trials<br>Sequencing for health facility-based and community-based research projects |
| Zambia | 0.27% (0.42%) | Yes | Yes | Yes | Yes | Not all investigations are being performed at all times. When cases exceed a particular threshold cluster, random and imported case surveillance reduces or stops. Total cases 8/2/21 = 63.573, 8/3/21 = 82,421. |

| | | | | | | |
|---|---|---|---|---|---|---|
| Democratic Republic of Congo (DRC) | 1.4 % (2.87%) | Yes | No | Yes | No | Regular surveillance is based on samples availability; the surveillance of imported cases is based on samples of travellers coming in DRC. there are also "sequencing based on a research project focused on respiratory infections (Andemia) |
| South Africa (FS) | | Yes | No | No | No | All samples with Cts lower that 30 are stored (with storage record). From 5 districts samples are selected randomly on a week basis (10 - 30) per district. From the ~15 000 stored samples no repeat testing has been identified within less than 90 days. |
| Ghana (Uhas) | 0.36% (0.12%) | Yes | Yes | No | No | Random surveillance based on clusters of cases. During periods of suspected widespread infections, cases are randomly selected and sequenced. |
| Tunisia | 0.04% | Yes | No | No | Yes | Random surveillance. Cases are randomly selected and sequenced. Some suspected reinfection cases are now tested in Sfax (Tunisia). |

| | | | | | | |
|---|---|---|---|---|---|---|
| Morocco | | Yes | Yes | Yes | Yes | Sequencing of 10% of Sample that are positif for S drop real time PCR test using (taqPath kit from thermo) . Sanger Sequencing of the entire S gene for the confirmation of mutation related to new varriants. WGS for the genomic surveillance over time et geographical localtion. |
| Equatorial Guinea | 3.10% | Yes | YES | Yes | No | During the first wave from March to August, all positive samples were stored and a random selection of these samples were sequenced. |
| Côte d'Ivoire (Bouaké) | 24.30% | Yes | No | No | No | Data set includes all CoV-2 RT-PCR samples tested positive from surveillance in regions of Côte d'Ivoire other than Abidjan; testing at CHU Bouaké; sampling period May-November 2020. Currently generating sequences from samples collected between Dec 2020 and March 2021. Calculation of cases (collumn C): suspected cases: 1199; of those tested: 100%; of those tested positive: 268 (22.36%); of those sequenced: 65 |

| | | | | | | |
|---|---|---|---|---|---|---|
| Algeria | 0,08% | Yes | Yes | Yes | No | Sequencing of Sample that are negatif for S by rRTPCR test using (taqPath kit from thermo) . Sanger Sequencing of the entire S gene for the confirmation of mutation related to new varriants. WGS for the genomic surveillance using MinION nanopore is in progress. |
| Mayotte | | Yes | Yes | No | No | Random surveillance, with extra samples collections in case of |
